# Use of Biochemical Biomarkers to Evaluate the Role of Oxidative Stress in the Progression of COVID-19 Severity

**DOI:** 10.1101/2024.08.11.24311803

**Authors:** Luis Marcano-Marcano, Raquel Salazar-Lugo

## Abstract

The role of oxidative stress and the use of biochemical biomarkers in the severity of COVID-19 was evaluated through a literature review (2020–2021) using scientific search engines such as PubMed, Science Direct, and Google Scholar. The search was limited to articles published in Spanish or English that reported on COVID-19 and its relationship with oxidative stress, following PRISMA-2020 guidelines. The search terms included oxidative stress, COVID-19, SARS-CoV-2, oxidative biomarkers, and oxidative damage. 93.5% of the selected studies were from the year 2021. These studies evaluated both oxidative stress biomarkers and oxidative damage biomarkers in COVID-19 patients. The reviewed studies reinforce the strong association of SARS-CoV-2 with oxidative stress and demonstrate how SARS-CoV-2-induced ROS production and disruption of the antioxidant defense system trigger a pro-inflammatory environment and cause severe tissue damage. In 64.7% of the studies, a combination of oxidative stress biomarkers (antioxidant and oxidative damage biomarkers) was used to assess COVID-19 severity. The most commonly used antioxidant biomarkers were thiols and total antioxidant capacity, followed by glutathione. The most commonly used oxidative damage biomarkers were malondialdehyde and peroxides, followed by advanced oxidation protein products. COVID-19 leads to a decrease in the antioxidant defense system, reflected by a decrease in antioxidant biomarkers and an increase in oxidative damage biomarkers.

## Introduction

The severe viral infection caused by COVID-19 has become a challenge for the scientific community due to the complexity of the biochemical and cellular events involved that drive the host’s pathological responses, contributing to the severity of the disease (Delgado-Roche and Mesta 2020). One of the aspects to consider in determining the severity of COVID-19 is the oxidative state presented by individuals, which could be a determinant of the infection’s evolution.

In general terms, respiratory viral infections are associated with inflammatory processes that may be linked to a redox imbalance or oxidative stress (Suhail et al. 2020). Based on an exhaustive literature analysis, Polonikov (2020) has proposed that endogenous glutathione deficiency, a tripeptide considered the main cellular antioxidant, could be a crucial factor in the increased oxidative damage induced by SARS-CoV-2 in the lungs. Consistent with these findings, it has been observed that the reduction of protein sulfhydryl groups (cysteine SH groups) weakens the binding of the SARS-CoV/CoV-2 spike protein, which points to a molecular basis that explains the severity of COVID-19 infection due to oxidative stress (Hati and Bhattacharyya 2020).

Oxidative stress (OS) is produced by an inadequate distribution between the production of reactive oxygen species (ROS) and antioxidant defenses (Betteridge 2000). Most scientists define OS as a vast region of reactions and interactions between ROS and other highly reactive species, including free radicals. The imbalance in the body’s antioxidant system produced by OS can be a life-threatening factor, as the overproduction of ROS and the deprivation of antioxidant mechanisms are crucial for viral replication and the subsequent disease associated with the SARS-CoV-2 virus. OS can occur or be controlled by many agents: genetic background, biological enzymatic processes, lifestyle, and all related aspects (Demirci-Çekiç et al., 2022). The close connection between respiratory viral infections and the generation and spread of ROS strongly involves OS in the severity of COVID-19 (Muhoberac 2020).

There are OS biomarkers that could be effectively used for the prognosis and monitoring of COVID-19. These biomarkers include antioxidant molecules (antioxidant vitamins; vitamin A, vitamin C, vitamin D, and vitamin E), molecules such as glutathione, thiols, and beta-carotene; antioxidant enzymes (endogenous glutathione (GSH), glutathione peroxidase (GPx), superoxide dismutase (SOD), free elements (Mn, Zn, Cu, Cr, and Se); and oxidative damage biomarkers (malondialdehyde, advanced oxidation protein products, ischemia-modified albumin, oxidized low-density lipoproteins), among others that stand out as oxidative damage biomarkers of lipids (Delgado-Roche and Mesta 2020), proteins (ischemia-modified albumin, advanced oxidation protein products (Cristani et al. 2015; Shevtsova et al. 202). Analytical follow-up of the antioxidant system can help prevent disease progression; thus, these biomarkers could be included in the prescribed tests to monitor COVID-19 patients, providing more analytical information to the physician.

As of February 1, 2022, the pandemic caused by the SARS-CoV-2 coronavirus, responsible for COVID-19, has resulted in 56 million deaths and over 376 million confirmed cases worldwide, with the United States of America leading in the number of infections and deaths, followed by India, Brazil, and Russia (WHO 2022). In Guatemala, by the end of January 2022, approximately 1% of the population (689,609 people) had been infected with the virus, with 16,379 deaths reported (WHO 2022).

This study aims to evaluate the role of oxidative stress in the progression of COVID-19 severity through the use of biochemical biomarkers; this will be done through a scoping review. Given the current relevance of this disease, it is important for the research group to compile information. The results of this scoping review will provide valuable information to clinical researchers and students in health-related fields interested in deepening their knowledge about the COVID-19 pandemic from a perspective that involves the individual’s OS as a major comorbidity for the progression of this disease.

## Materials and Methods

### Study Design

This work was conducted as a scoping review. A scoping review aims to extract key concepts from cutting-edge literature in a research area. This design was chosen because it provides a broader scope of specific field coverage. In this particular review, the purpose is to clarify concepts and inform the state of the art regarding oxidative stress and its relationship with COVID-19 severity, as well as to report on the analytical tests used to evaluate the oxidative status of individuals with COVID-19. The inclusion criteria applied were: articles in Spanish or English, experimental or clinical, published in the last 2 years (due to the novelty of the topic), that included the full text, and that focused on assessing oxidative stress as a risk factor for the development of COVID-19 severity.

The study was conducted in Guatemala City from February 2020 to January 2022.

### Instruments and Data Analysis

This scoping review followed the steps of a systematic review but with a broader scope. PRISMA guidelines for systematic reviews and the Joanna Briggs Institute guidelines for scoping reviews were adhered to (Peters, 2015).

For data collection, the procedure proposed by Arksey and O’Malley (2007) was used. For the development of this scoping review, original articles published in the last two years (2020–October 2021) were included from the following databases: PubMed, Google Scholar, and ScienceDirect. The search was limited to full-text articles published in Spanish or English and focused on original research reporting on COVID-19 and its relationship with oxidative stress according to the PRISMA-2009 checklist criteria.

The following search terms were used: oxidative stress, COVID-19, oxidative damage, SARS-CoV-2, oxidative stress biomarkers, and oxidative damage biomarkers. The search terms were kept broad to cover all potential applicable studies.

## Results

A total of 1,536 abstracts were identified, and 65 documents were categorized after primary exclusion. Of the categorized works, 23 studies from indexed scientific journals were included (Figure 1).

**Figure 1.**
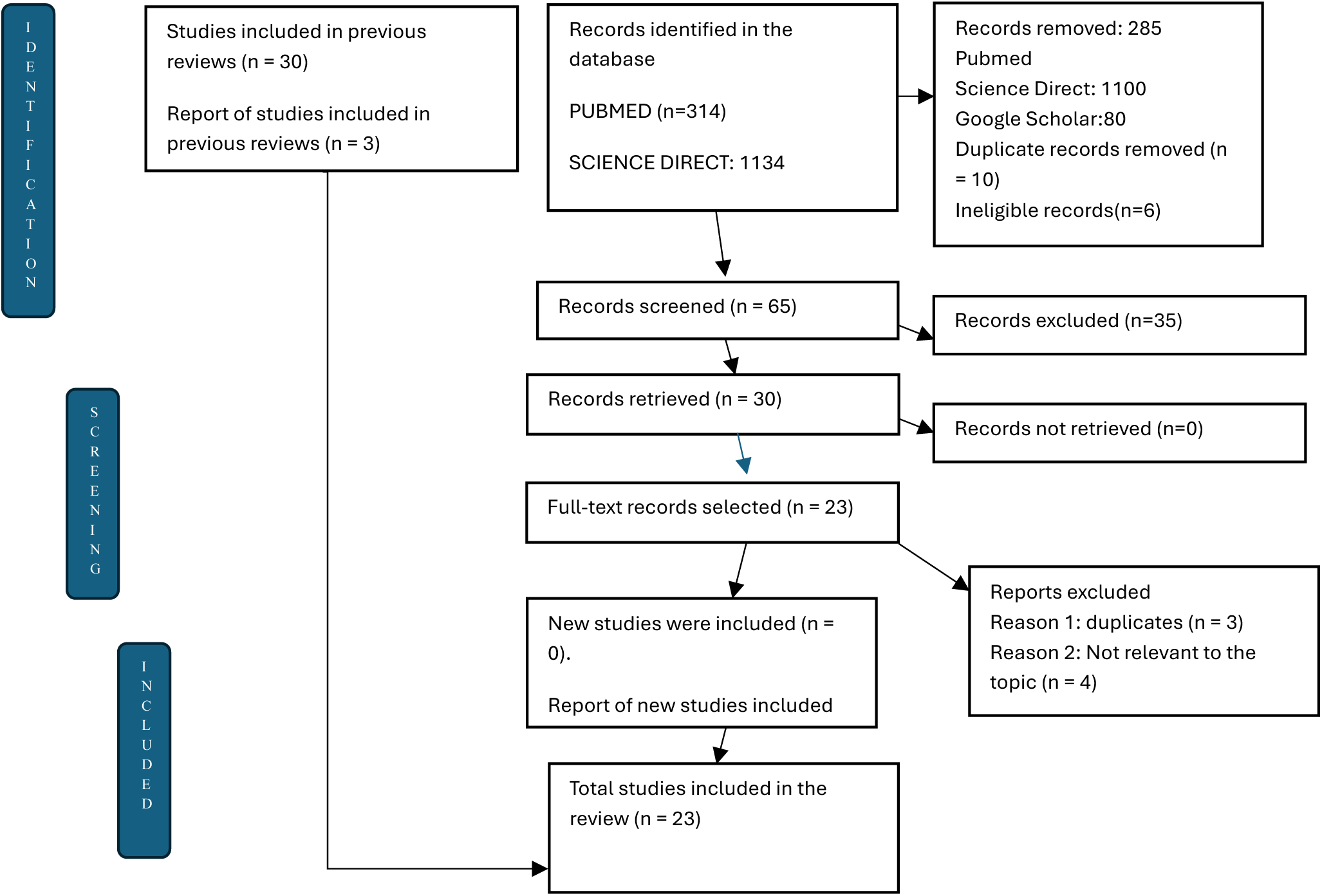
PRISMA 2020 flow diagram for updated scoping review, including database research (Page et al., 2021).

93.5% of the selected studies are from the year 2021. Six (6) studies were basic research investigations whose results show evidence linking the redox state of the cell with COVID-19 (Table 1). The studies present results indicating that an oxidized state of the molecules involved in viral recognition and cell entry facilitates infection, as indicated by Hati and Bhattacharyya (2021) in a study based on a simulation of the proteins involved in the initial viral recognition, such as the ACE2 receptor and the viral spike protein, and their conformational arrangement in an oxidized state, which allows greater success in viral recognition and thus infection.

**Table 1.**
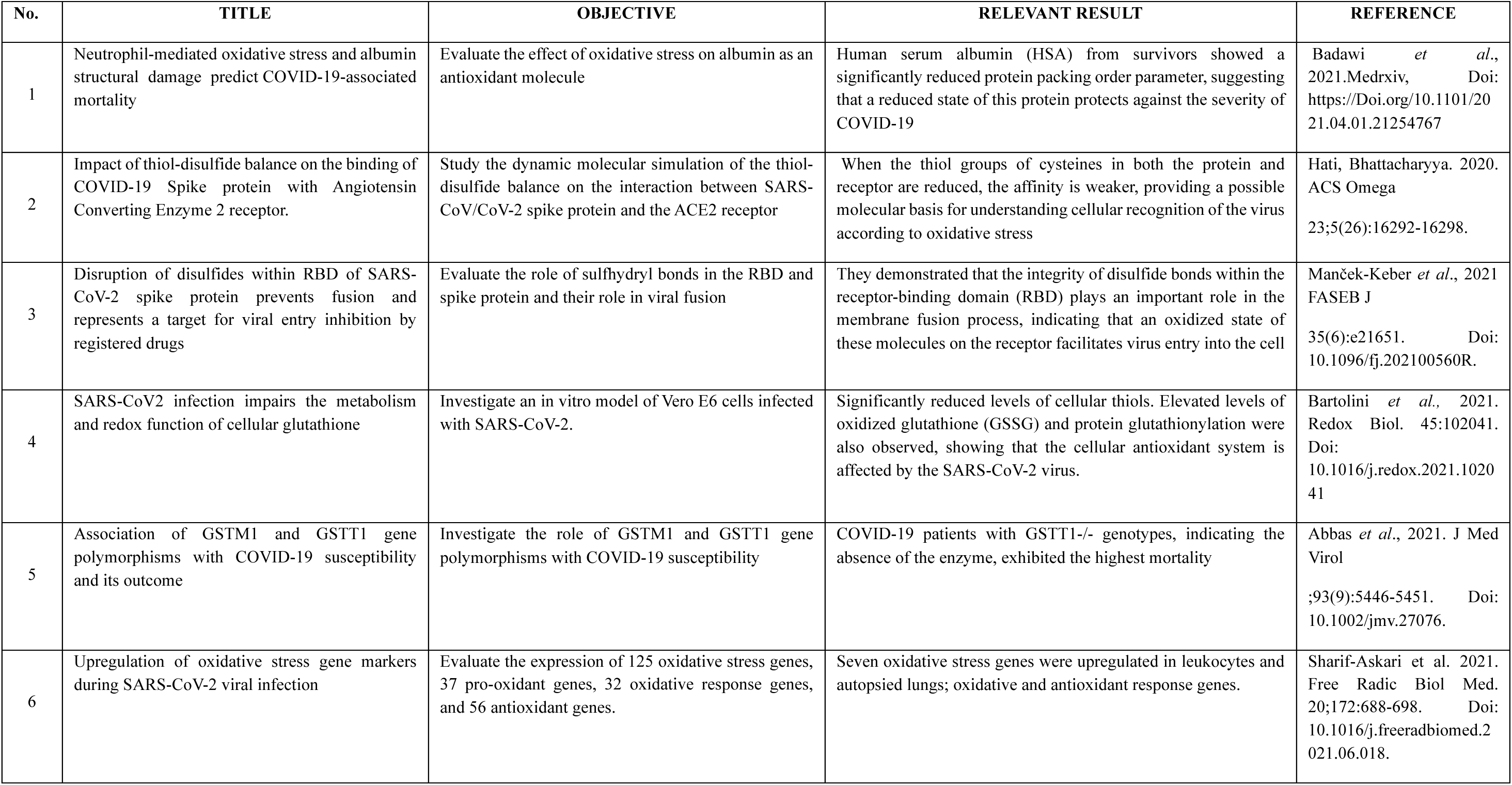
Experimental basic research studies evaluating the role of the oxidative system in the progression of COVID-19.

Manek-Keber et al. (2021) experimentally demonstrate the role of the oxidized state of the ACE2 receptor and the spike protein in the success of SARS-CoV-2 infection, coinciding with what was proposed by Hati and Bhattacharyya (2020) in their molecular simulation study.

Another study conducted on Vero 6 cells, which the authors identify as a well-characterized in vitro model of SARS-CoV-2 infection, showed the involvement of the organism’s oxidative system in infection progression (Bartolini et al., 2021).

Studies on proteins directly related to oxidative stress showed that COVID-19 patients with the GSTT1-/-genotype, which codes for the glutathione S-transferase (GST) enzyme, exhibited higher mortality (Abbas et al., 2021). This is an important enzyme that catalyzes the conjugation of glutathione (GSH) with electrophiles to protect the cell from oxidative damage and participates in the antioxidant defense mechanism in the lungs. It was also demonstrated that oxidative and antioxidant response genes were upregulated in the lungs and leukocytes of COVID-19 patients (Sharif-Askari et al., 2021).

The aforementioned findings constitute evidence of the involvement of the organism’s oxidative state in the progression of the disease caused by the SARS-CoV-2 virus and indicate that an oxidized environment facilitates the virulence and severity of SARS-CoV-2. This may explain why the severity of infection is greater in people with underlying pathologies associated with reduced antioxidant defenses (Korakas et al., 2020; Cordtz et al., 2021).

The second group of selected works responded to the keywords COVID-19/oxidative stress and the established criterion that they were not reviews, case studies, editorials, hypotheses, or comments. Seventeen studies were selected, of which 11 were clinical research studies, one was a pilot study, one was a cross-sectional comparative study, one was a single-center retrospective study, one was an observational study, and one was a multicenter study (Table 2).

**Table 2.**
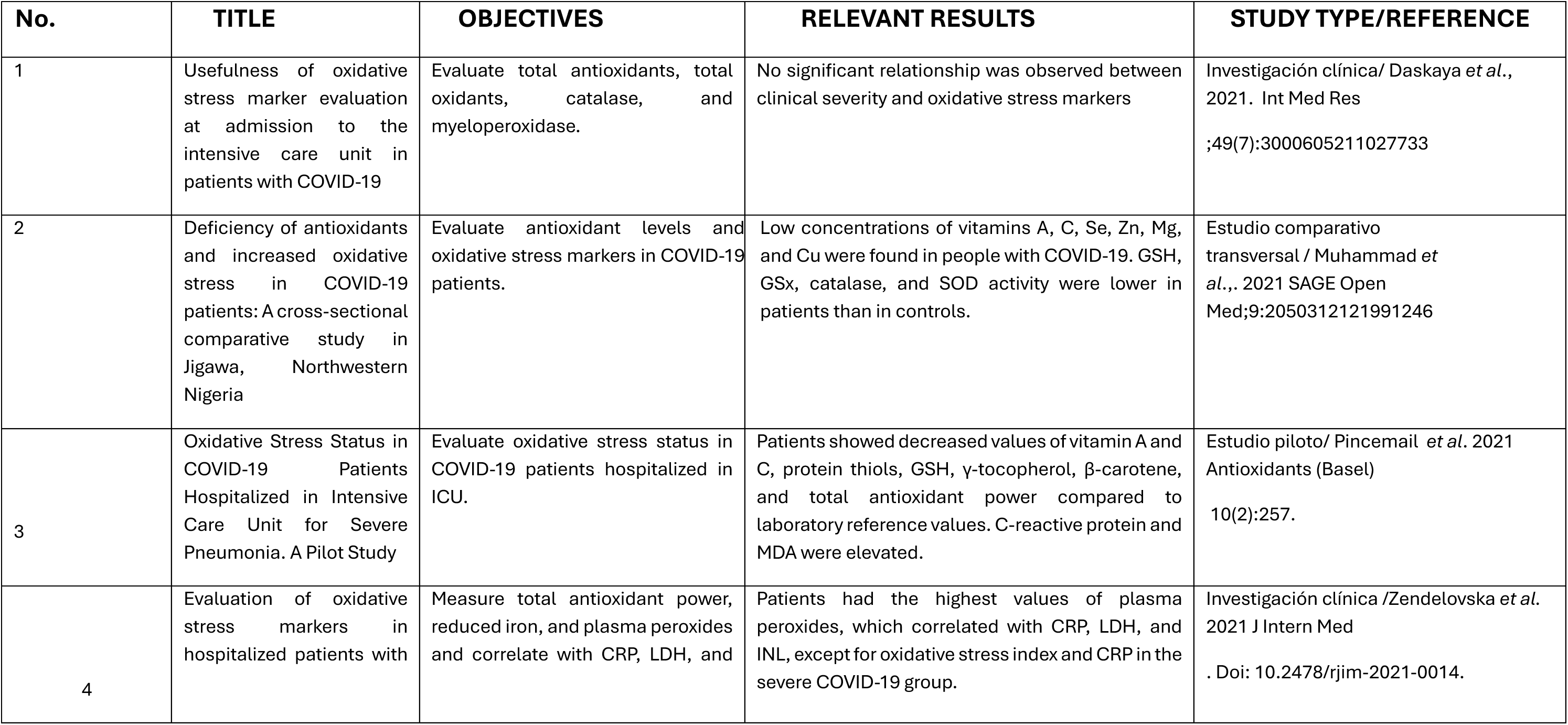
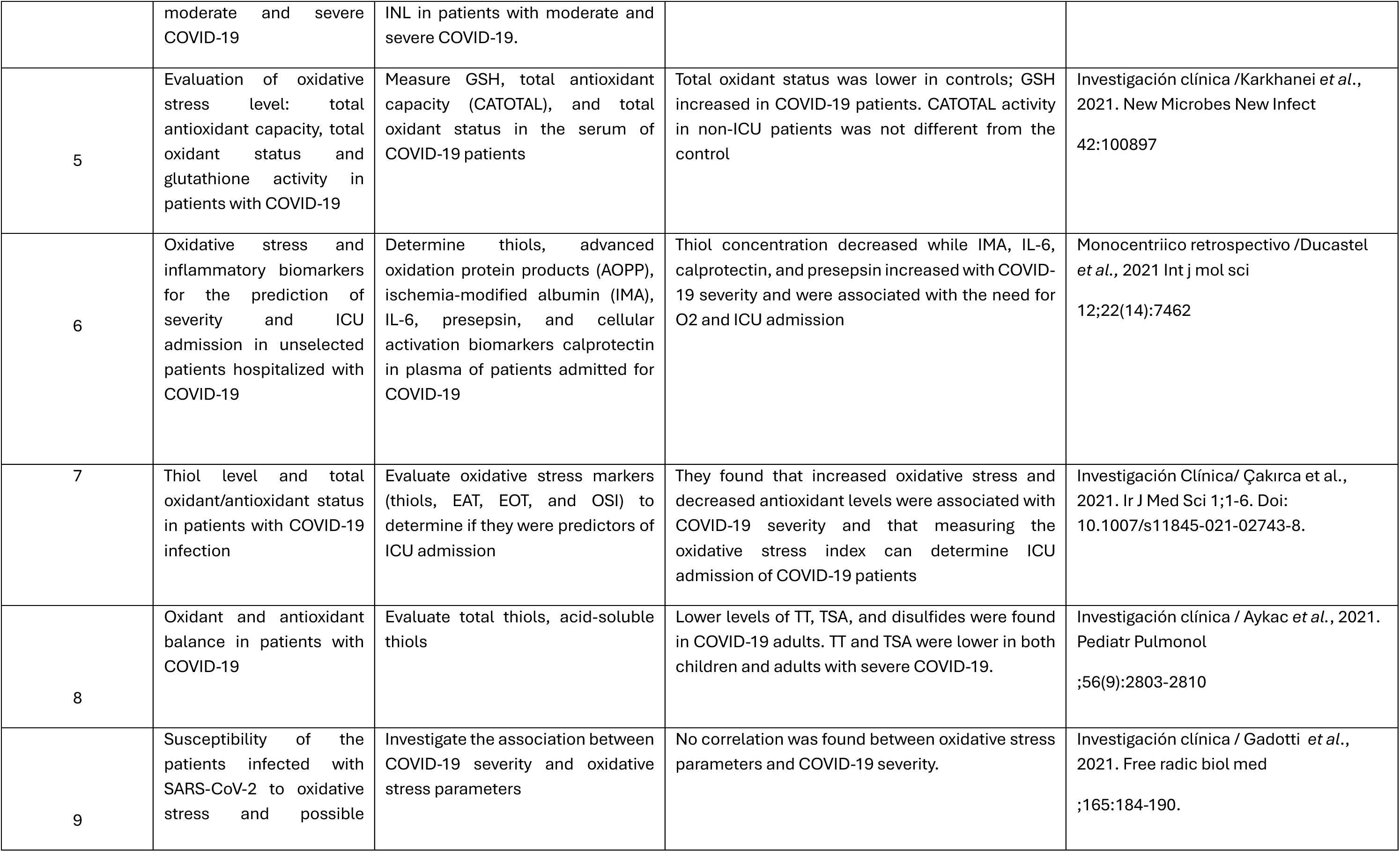
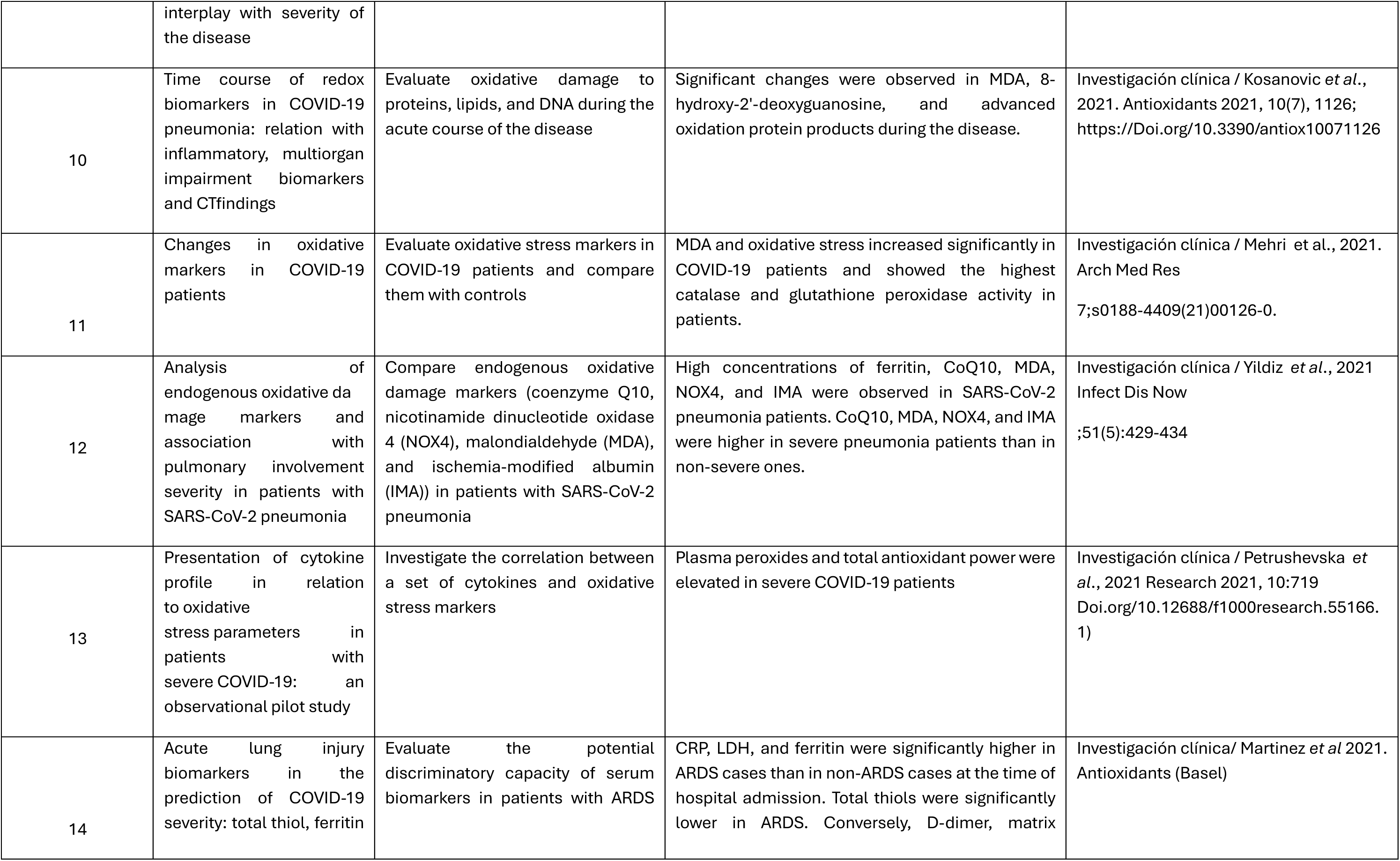
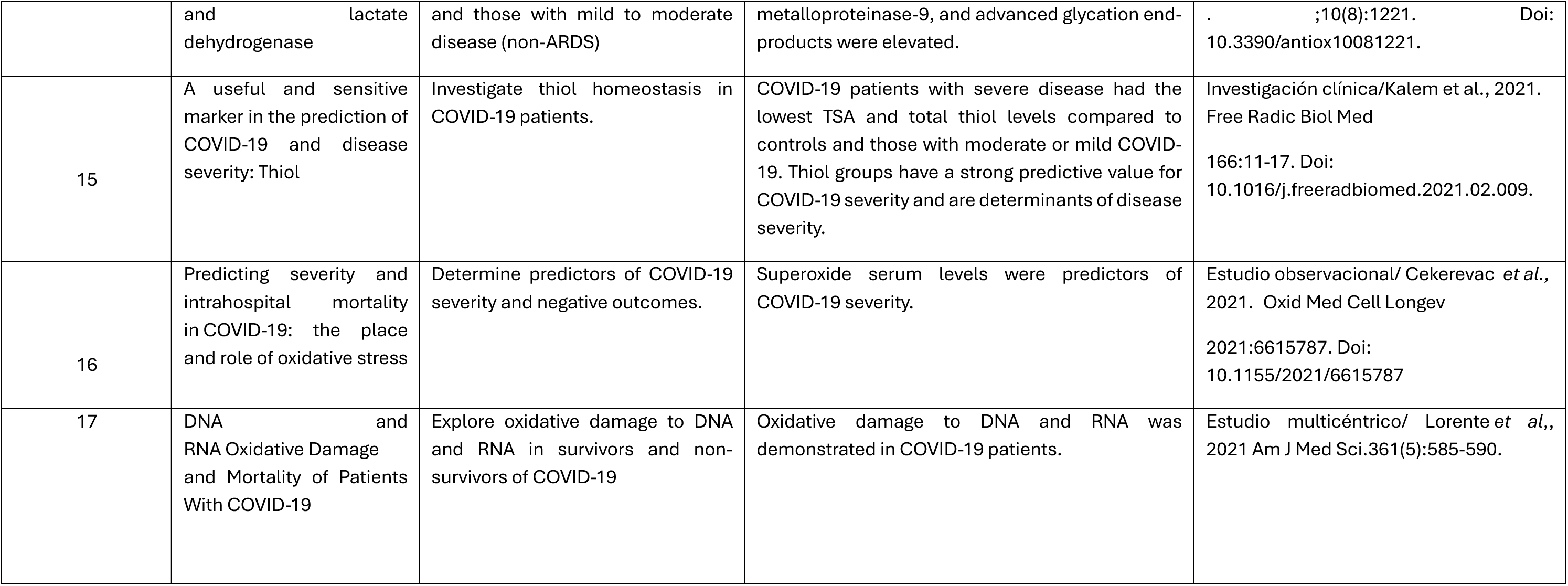
Studies evaluating oxidative stress based on primary metabolites of the antioxidant system generated or participating in the oxidative stress process in COVID-19 patients.

The results obtained in seven (7) of the reviewed studies evaluated thiols as an indicator of the oxidative state of COVID-19 patients, concluding that these molecules are predictors of severity and admission to the ICU (intensive care unit) for COVID-19 patients (Ducastel et al. 2021; Pincemail et al. 2021; Çakırca et al. 2021; Aykac et al. 2021; Martinez et al. 2021; Kalem et al. 2021; Cekerevac et al. 2021; Table 2).

In studies where glutathione concentrations were determined, it was found that this parameter was decreased in COVID-19 patients (Muhammad et al., 2021; Pincemail et al., 2021; Karkhanei et al., 2021; Gadotti et al., 2021; Cekerevac et al., 2021; Table 2).

COVID-19 patients showed an increase in the activity of antioxidant enzymes such as superoxide dismutase, catalase, and glutathione peroxidase (Pincemail et al., 2021; Muhammad et al., 2021; Gadotti et al., 2021; Mehri et al., 2021; Cekerevac et al., 2021). Oxidative stress biomarkers belonging to the category of oxidative damage indicators, such as malondialdehyde (MDA), ischemia-modified albumin (IMA), sulfhydryl groups, advanced glycation end products (AGEs), oxidized guanine, and peroxides, were elevated in COVID-19 patients and increased with the severity of the disease (Daskaya et al. 2021; Muhammad et al. 2021; Pincemail et al. 2021; Zendelovska et al. 2021; Ducastel et al. 2021; Aykac et al. 2021; Gadotti et al. 2021; Cekerevac et al. 2021; Kosanovic et al. 2021; Mehri et al. 2021; Yildiz et al. 2021; Petrushevska et al. 2021; Martinez et al. 2021; Lorente et al. 2021; Table 2).

The results reported regarding the oxidative stress biomarkers evaluated in COVID-19 patients indicate the high commitment of the body’s antioxidant system to the pathogenesis of the disease caused by the SARS-CoV-2 virus (Table 2).

In Table 3, the two categories of oxidative stress biomarkers evaluated in the reviewed studies are specified. In 11 of the 17 (64.7%) reviewed studies, a combination of antioxidant biomarkers and oxidative damage biomarkers was used.

**Table 3.** Oxidative stress biomarkers evaluated in COVID-19 patients.

| No. | TITLE | EVALUED OXIDATIVE STRESS BIOMARKERS |  | REFERENCE |
| --- | --- | --- | --- | --- |
|  |  | EVALUED OXIDATIVE STRESS BIOMARKERS | ANTIOXIDANT BIOMARKERS |  |
| 1 | Usefulness of oxidative stress marker evaluation at admission to the intensive care unit in patients with COVID-19 | Total antioxidant status, total oxidant status, catalase. | Ferritin, myeloperoxidase | Daskaya *et al.*, 2021. Int Med Res<br>49(7):3000605211027733 |
| 2 | Deficiency of antioxidants and increased oxidative stress in COVID-19 patients: A cross-sectional comparative study in Jigawa, Northwestern Nigeria | Glutathione, superoxide dismutase, glutathione peroxidase, vitamins A and D | Malondialdehyde, 8-isoprostaglandin F2 alpha | Muhammad *et al.*, 2021 SAGE Open Med;9:2050312121991246 |
| 3 | Oxidative Stress Status in COVID-19 Patients Hospitalized in Intensive Care Unit for Severe Pneumonia. A Pilot Study | Glutathione, protein thiols, glutathione peroxidase, vitamin C and E, beta-carotene, selenium, zinc, and copper, total antioxidant capacity | Lipid peroxides, myeloperoxidase, oxidized LDL | Pincemail *et al.* 2021 Antioxidants (Basel)<br>10(2):257. |
| 4 | Evaluation of oxidative stress markers in hospitalized patients with moderate and severe COVID-19 | Total antioxidant power | Plasma peroxides | Zendelovska *et al.* 2021 J Intern Med<br>Doi: 10.2478/rjim-2021-0014. |
| 5 | Evaluation of oxidative stress level: total antioxidant capacity, total oxidant status and glutathione activity in patients with COVID-19 | Glutathione, total antioxidant capacity | Not evaluated | Karkhanei *et al.*, 2021. New Microbes New Infect ;42:100897 |
| 6 | Oxidative stress and inflammatory biomarkers for the prediction of severity and ICU admission in unselected patients hospitalized with COVID-19 | Total thiols | Advanced oxidation protein products, ischemia-modified albumin | Ducastel *et al.*, 2021 Int J Mol Sci 12;22(14):7462 |
| 7 | Thiol level and total oxidant/antioxidant status in patients with COVID-19 infection | Total thiols, total antioxidant status, total oxidative status, oxidative stress index | Not evaluated | Çakırca *et al.*, 2021. Ir J Med Sci 1;1-6. Doi: 10.1007/s11845-021-02743-8. |
| 8 | Oxidant and antioxidant balance in patients with COVID-19 | Total thiols, acid-soluble thiols | Sulfhydryl groups | Investigación clínica / Aykac *et al.*, 2021. Pediatr Pulmonol 56(9):2803-2810 |
| 9 | Susceptibility of the patients infected with SARS-CoV-2 to oxidative stress and possible interplay with severity of the disease | Glutathione, total antioxidant capacity, glutathione transferase | Malondialdehyde, protein carbonylation, sulfhydryl groups, peroxides | Gadotti *et al.*, 2021. Free radic biol med ;165:184-190. |
|  | Time course of redox biomarkers in COVID-19 pneumonia: relation with inflammatory, multiorgan | Not evaluated | Malondialdehyde, 8-hydroxy-2'-deoxyguanosine, and advanced oxidation protein products | Kosanovic *et al.*, 2021. Antioxidants 10(7), 1126; |
| 10 | impairment biomarkers and CT findings |  |  | https://doi.org/10.3390/antiox10071126 |
| 11 | The comparison of oxidative markers between COVID-19 patients and healthy subjects | Catalase, superoxide dismutase, total oxidative status | Malondialdehyde | Mehri et al., 2021. Arch Med Res 7;s0188-4409(21)00126-0. |
| 12 | Analysis of endogenous oxidative damage markers and association with pulmonary involvement severity in patients with SARS-CoV-2 pneumonia | Not evaluated | Ferritin, malondialdehyde, ischemia-modified albumin, nicotinamide adenine dinucleotide oxidase 4 (NOX4), coenzyme Q10 | Yildiz et al., 2021 Infect Dis Now 51(5):429-434 |
| 13 | Presentation of cytokine profile in relation to oxidative stress parameters in patients with severe COVID-19: an observational pilot study | Total antioxidant power, oxidative stress index | Plasma peroxides | Petrushevska et al., 2021 Research 10:719<br>[Doi.org/10.12688/f1000research.55166.1](https://doi.org/10.12688/f1000research.55166.1) |
| 14 | Acute lung injury biomarkers in the prediction of COVID-19 severity: total thiol, ferritin and lactate dehydrogenase | Total thiols | Matrix metalloproteinase-9, advanced glycation end products, ferritin | Martinez et al 2021. Antioxidants (Basel). ;10(8):1221. Doi: 10.3390/antiox10081221. |
| 15 | A useful and sensitive marker in the prediction of COVID-19 and disease severity: Thiol | Total thiols | Not evaluated | Kalem *et al.*, 2021. Free Radic Biol Med<br>;166:11-17.Doi:<br>10.1016/j.freeradbiomed.2021.02.009. |
| 16 | Predicting severity and intrahospital mortality in COVID-19: the place and role of oxidative stress | Glutathione, catalase, and superoxide dismutase | Lipid peroxidation index (TBARS), peroxides, and superoxide | Cekerevac *et al.*, 2021. Oxid Med Cell Longev<br>;2021:6615787. Doi:<br>10.1155/2021/6615787 |
| 17 | DNA and RNA Oxidative Damage and Mortality of Patients With COVID-19 | Not evaluated | Oxidized guanine | Lorente *et al.*, 2021 Am J Med Sci<br>.361(5):585-590. |

Among the antioxidant biomarkers, the most commonly used were thiols and total antioxidant power, followed by the determination of glutathione, the measurement of antioxidant enzyme activity, and, to a lesser extent, the determination of selenium and zinc, beta-carotene, and vitamins A, D, C, and E (Figure 2).

**Figure 2.**
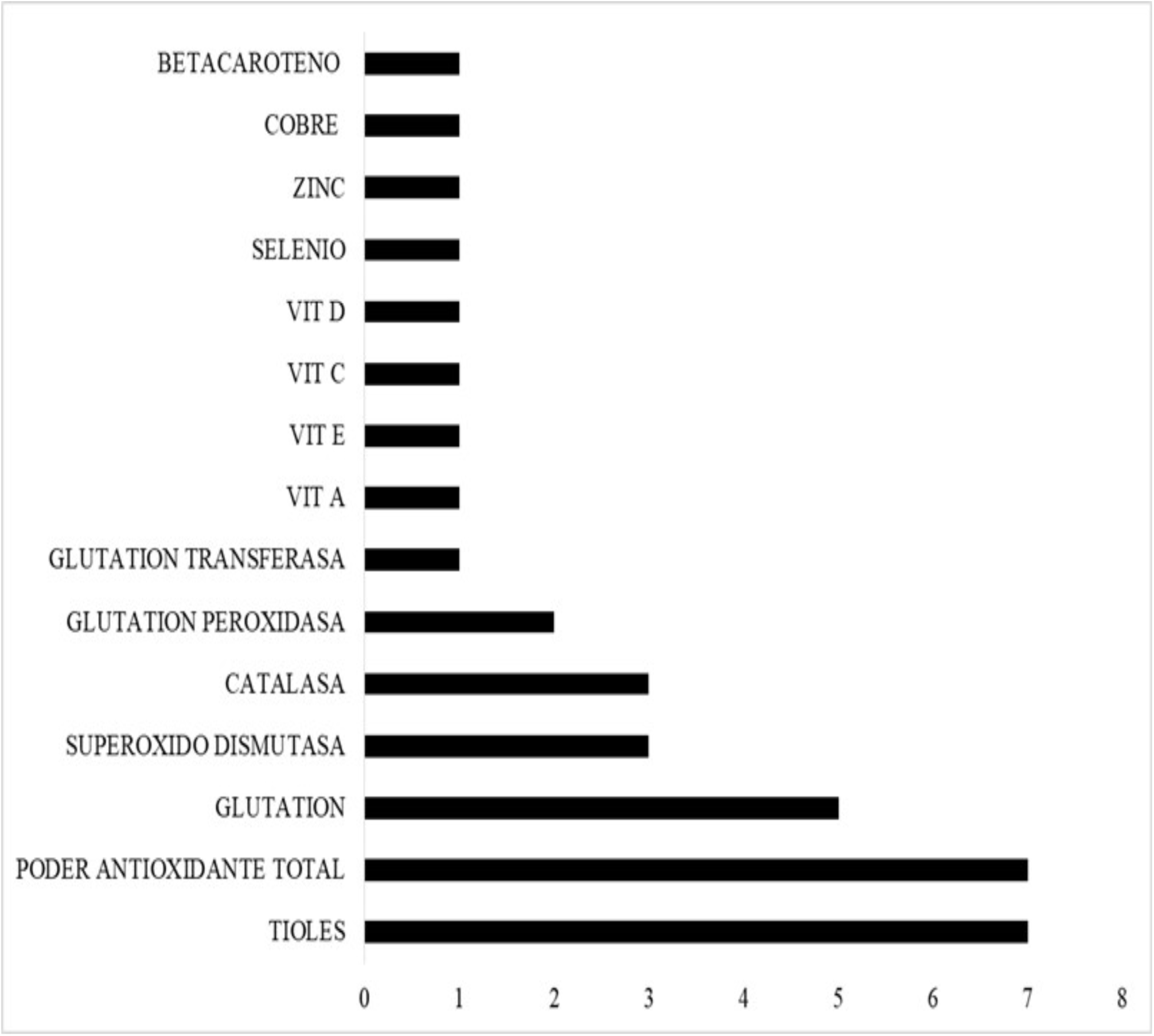
Antioxidant biomarkers determined in COVID-19 patients according to the reviewed literature. Years 2020-2021. Abbreviations: Vit D: vitamin D; Vit C: vitamin C; Vit E: vitamin E; Vit A: vitamin A.

On the other hand, the most commonly employed oxidative damage biomarkers were the determination of malondialdehyde and peroxides, followed by the determination of advanced oxidation protein products, ferritin, and, to a lesser extent, other indicators such as the measurement of myeloperoxidase enzyme activity, the quantification of ischemia-modified albumin, and the determination of lipid and plasma peroxides and oxidized guanine (Figure 3).

**Figure 3.**
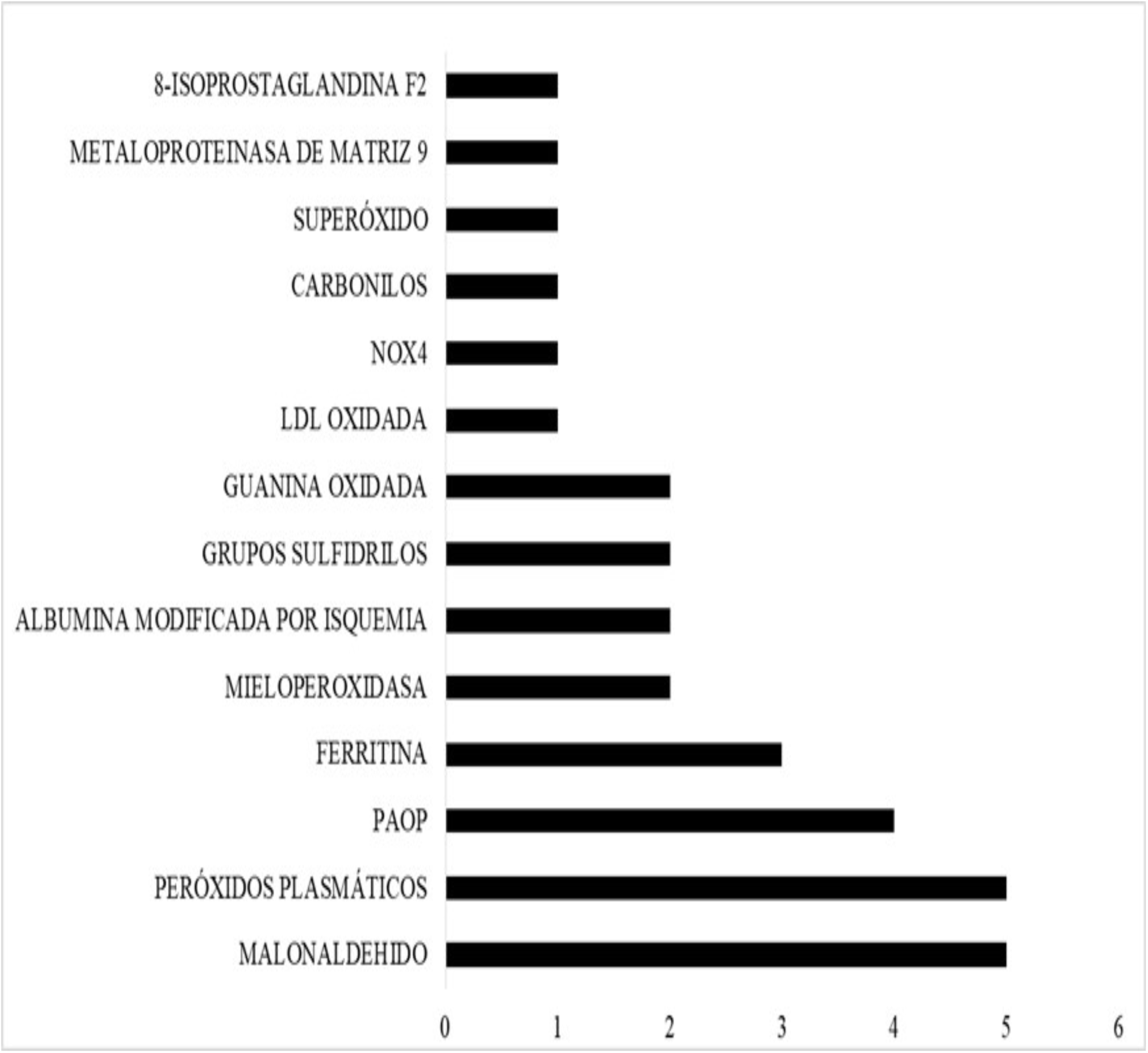
Oxidative damage biomarkers determined in COVID-19 patients according to the reviewed literature. Years 2020-2021. Abbreviations: PAOP: Advanced oxidation protein products; NOX4: Nicotinamide adenine dinucleotide oxidase 4.

## Discussion

This scoping review synthesizes the emerging global evidence on the association of oxidative stress with the progression of COVID-19 pathology and how this can explain the severity of this viral infection. The reviewed studies reinforce what was initially noted at the beginning of the COVID-19 pandemic by some authors about the central role of oxidative stress in the pathogenesis of this disease caused by the SARS-CoV-2 coronavirus. The reviewed literature indicates that the individual’s redox state is diminished in the pathophysiological process of this disease.

The reviewed experimental studies provide evidence of the connection between the oxidative state of the body and the virulence and progression of COVID-19, as demonstrated by Abbas et al. (2021), who found that patients with the GSTT1-/-genotype showed the highest mortality from COVID-19. Similarly, Sharif-Askari et al. (2021) demonstrated that the SARS-CoV-2 virus induces the expression of oxidative stress genes. Manek-Keber et al. (2021) highlight the importance of disulfide bonds in the binding proteins of the virus to the receptor in the success of fusion and virulence, and Bartolini et al. (2021) demonstrate in Vero cells that the cellular antioxidant system is affected by the SARS-CoV-2 virus.

Regarding the oxidative stress biomarkers evaluated in COVID-19 patients, the reviewed studies show that a variety of biomarkers are being used. The determination of thiol groups, including total thiols (TT), acid-soluble thiols (TSA), and total antioxidant power, was the most commonly used. Some authors suggest that the determination of thiol groups is a sensitive biomarker to evaluate the oxidative state in COVID-19 patients (Pincemail et al., 2021; Ducastel et al., 2021; Çakırca et al., 2021; Aykac et al., 2021; Martinez et al., 2021; Kalem et al., 2021). Kalem et al. (2021) found a negative correlation between symptoms and TT and TSA levels in COVID-19 patients, indicating that there is a consumption of antioxidant molecules as the body fights the infection. Similarly, Ducastel et al. (2021) reported that a thiol concentration of <154 µmol/l was predictive of ICU admission in COVID-19 patients.

Thiol groups, which are predominantly found in protein structures, can form reversible disulfide bonds due to the presence of oxidants, maintaining a dynamic homeostasis of thiols and sulfhydryl groups. Since they are found in proteins, these groups are the body’s major antioxidants and play a crucial role against oxidative stress and in protecting against damage induced by free radicals. They are present both in cells and in plasma and serve as primary regulators of oxidative stress. In cases where there are high levels of oxidative stress, thiol levels decrease because they participate in neutralizing free radicals (Ulrich and Jakob 2019).

The determination of glutathione represented the second most evaluated oxidative stress biomarker in COVID-19 patients (Muhammad et al., 2021; Pincemail et al., 2021; Karkhanei et al., 2021; Gadotti et al., 2021; Cekerevac et al., 2021). Glutathione is a crucial antioxidant, well known for modulating the behavior of many cells, including immune system cells, enhancing innate and adaptive immunity, and providing protection against microbial, viral, and parasitic infections. According to Forman et al. (2016), low levels of GSH are associated with abnormalities in the pulmonary surfactant system, while normal intracellular GSH levels can exert critical negative control over the elaboration of pro-inflammatory cytokines.

Oxidative damage biomarkers such as malondialdehyde (MDA), an indicator of lipid peroxidation; 8-hydroxy-2’-deoxyguanosine (8-OHdG), a marker of DNA damage; ischemia-modified albumin (IMA); and advanced oxidation protein products (AOPP), which are biomarkers of protein structure damage, were also determined by some authors (Kosanovic et al., 2021; Yildiz et al., 2021; Ducastel et al., 2021; Daskaya et al., 2021).

Among the biomarkers mentioned in the previous paragraph, malondialdehyde, followed by the determination of advanced oxidation protein products, was the most frequently used. Oxidative damage biomarkers varied with the course of the disease and correlated with some inflammatory and tissue damage biomarkers (Kosanovic et al., 2021). Merhi et al. (2021) found that malondialdehyde levels in the serum of patients were significantly higher in the ICU group compared to non-ICU COVID-19 patients.

The production of free radicals during SARS-CoV-2 infection has the ability to modify macromolecules, causing damage to them. Oxidative damage to DNA and RNA has been reported in severe COVID-19 patients admitted to the ICU (Lorente et al., 2021).

The reviewed studies reinforce the strong association of SARS-CoV-2 with oxidative stress (OS) that triggers cytokine production, inflammation, and other pathophysiological activities (Li et al. 2020). The production of ROS induced by SARS-CoV-2 disrupts the antioxidant defense system, which triggers a pro-inflammatory environment and severe tissue damage, contributing to fatal outcomes in patients. This would be one of the main reasons why patients with pre-existing conditions such as diabetes, hypertension, and pulmonary, cardiac, and renal diseases, which are pathologies associated with chronic inflammatory processes and oxidative imbalance, have a higher risk of developing severe infection (Lima-Martínez et al., 2021; Mihalopoulos et al., 2020; Li et al., 2020).

Similarly, the reviewed studies provide evidence of the well-known association between inflammation and oxidative stress. In the case of this global viral infection, the high production of free radicals by primary immune system cells such as neutrophils and macrophages (Borges et al. 2020) can oxidize macromolecules, as evidenced by the reviewed studies. This excessive production of ROS can structurally modify proteins, affect the metabolome, and modulate gene cascades involved in inflammatory response signaling pathways (Shi et al., 2021; Sharif-Askari et al., 2021), creating an oxidative-inflammatory spiral that characterizes severe cases of COVID-19 (Zendelovska et al., 2021), thus affecting all organs and facilitating the progression of the disease to more severe stages in patients with pre-existing conditions.

## Conclusions

The data provided in the reviewed experimental studies demonstrate the involvement of the oxidized state of the body in the success of SARS-CoV-2 infection and disease progression. This is evidenced by the significant decrease in antioxidant biomarkers in people with COVID-19, among them glutathione, thiols, total antioxidant capacity, antioxidant enzyme activity, and other antioxidant parameters such as beta-carotenes, zinc, selenium, and vitamins A, D, C, and E, as well as the significant increase in oxidative damage biomarkers such as malondialdehyde, advanced protein glycation end products, ferritin, ischemia-modified albumin, and lipid and plasma peroxides.

In 64.7% of the reviewed studies, a combination of oxidative stress biomarkers (antioxidant biomarkers) and oxidative damage was used. The most commonly used antioxidant biomarkers were thiol determination and total antioxidant power, followed by glutathione determination. The most commonly used oxidative damage biomarkers were malondialdehyde determination and peroxides, followed by advanced oxidation protein products.

## Strengths and limitations

This scoping review focuses on the relationship between COVID-19 and oxidative stress, mapping the different biomarkers being used to monitor the oxidative state of COVID-19 patients. The study focused on publications of clinical research as well as experimental studies that could provide evidence of the central role of oxidative stress in the progression of disease severity.

This review also has limitations; one of them is that only three databases were used for the review of selected studies, which may have excluded some relevant studies on the topic.

## Data Availability

All data produced in the present work are contained in the manuscript

## Acknowledgments

The authors would like to thank Licenciada Alejandra Sierra for her critical reading of the manuscript. We also extend our gratitude to Universidad Mariano Gálvez de Guatemala.

The authors declare that there are no conflicts of interest in the preparation of this work.

